# Thyroid volume, lobe asymmetry and AP diameter classification - A preliminary study

**DOI:** 10.1101/2025.01.11.25320363

**Authors:** Sanjida Akter, Azmal K Sarker, Shahriar Muttakin, Sanjoy Biswas

## Abstract

**Objective:** To assess the thyroid lobe volumes and the total thyroid volumes across the arbitrary range for AP diameter of thyroid lobes, the age and sex related variation and compare local normal sizes with those in the reference. Because assessment of thyroid gland size by visual comparison with anatomical relations or by an arbitrary category for antero-posterior (AP) diameter precludes use of management guidelines based on thyroid gland volume. Moreover, the international references for AP diameter seem larger for Bangladeshi patients and therefore deemed as inapplicable.

**Methods:** Patients who underwent thyroid ultrasound with thyroid volume measurements from October 2023 to October 2024 were included. The patients were classified into two groups. Group 1 consisted of children aged below 15 years while group 2 consisted of adolescents aged 15 years or more and all adults. Statistical analyses were applied to assess distribution of gland size between sexes and age groups, correlation between gland size and age, and distribution of thyroid lobe volume and total gland volume, across the arbitrary categories of AP diameter.

**Results:** Total 110 patients (90 females) with mean (± SD) age of 28.1 ± 13.2 were included. There was no significant difference of gland size between males and females. The AP diameter and right thyroid lobe was significantly larger than the left in group 2 consisting of adolescents and adults (p < 0.05). There was a positive correlation of thyroid size and age in group 1 consisting of pre-adolescents who had normal sized glands (R = 0.91, p < 0.01). Observation of thyroid volumes across the arbitrary AP diameter category showed AP diameter categories of right lobe tended to better classify the normal glands whereas the AP diameter categories of left lobe tended to perform better for classification of the enlarged glands.

**Conclusions:** This study apart from describing the interaction of age and sex with thyroid size, highlights the importance of considering the natural asymmetry between the thyroid lobes for accurate thyroid volume classification. This study points toward the void for a reference local standard of thyroid volume that can classify not only the different categories of enlargement but also the small volume glands.

## INTRODUCTION

Assessment of thyroid gland size, one of the major goals in the thyroid ultrasound, is often achieved by measuring the anteroposterior (AP) diameter of the individual lobes. The reference normal limits of AP diameters of the thyroid lobe show large variation and high upper limits (1-7), which barely match with the reported values from Bangladesh (8) and therefore are considered to be inapplicable for the patients in Bangladesh. The method of comparing anterior gland margin with the anterior margin of common carotid artery in the short axis view (3) gives an alternative non-quantitative understanding about thyroid enlargement to the sonographer. However, the quantitative estimation of thyroid enlargement is useful for goiter size determination before surgery (9-11), calculation of the dose of iodine-131 (12-14) and evaluate response to suppression treatments (15, 16). An arbitrary range for AP diameter of thyroid lobes has been in use for differentiating normal size from enlargement as well as for classification of thyroid enlargement into mild, moderate and marked categories. Although this AP diameter of thyroid lobes can give an idea about the gland size, the estimation of total gland volume would render the assessment of thyroid size useful for clinicians who incline to adhere to international guidelines of disease management (17).

The thyroid volume not only differs across the age and sex, but also has been observed to be affected by anthropometric indices, body composition proportions, serum levels of thyroid hormones and thyrotropin, environmental factors, physiological states and metabolic diseases. (18-40).

This retrospective study was done to understand the thyroid lobe volumes and the total thyroid volumes across the non-standard arbitrary range for AP diameter of thyroid lobes. Furthermore, the data was checked for the age and sex related variation of the thyroid volumes. A comparative evaluation of the normal AP diameter of thyroid lobes from the current study with those from the references was finally done.

## PATIENTS AND METHODS

All consecutive patients who underwent thyroid ultrasound with thyroid volume measurements from October 2023 to October 2024 were retrospectively included in this study. In all patients the volumes of both the right and left thyroid lobe were measured by biplanar method. Total gland volume was determined by adding up the volume of both right and lobe lobes. For the qualitative categorization of a lobe into normal or enlarged, an arbitrary limit for the AP diameter was used (Box 1) in addition to the criteria of comparing anterior gland margin with that of the common carotid artery in the short axis view, without manual compression. Patients whose thyroids were visually normal as well as their lobes’ AP diameter being within the arbitrary normal range (Box 1) were classified as normal sized gland (NSG).

Furthermore, the patients were classified into two groups. Group 1 consisted of children aged below 15 years while group 2 consisted of adolescents aged 15 years or more and all adults. The correlation between total gland volume and age was checked within the NSG patients.

To understand the distribution of thyroid lobe volume and total gland volume, across the arbitrary categories of AP diameter ranging from normal to three categories of enlargement, the mean volumes across the arbitrary AP diameter categories were checked.

All statistical tests were done on R. Correlation plots were generated using the “ggplot2” and the box-violin plots were generated using the “ggstatsplot” functions. The forest plot was generated using Microsoft Excel.

### Box 1.

Arbitrary limits for AP diameter categories

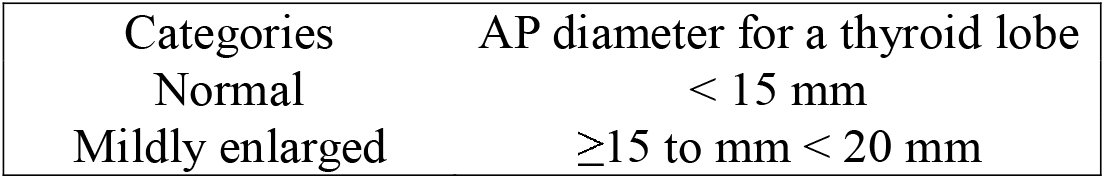
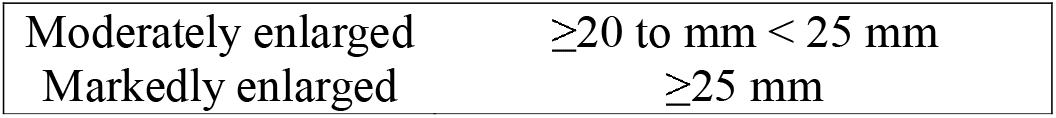

## RESULTS

A total of 110 patients (90 females) with mean (± SD) age of 28.1 ± 13.2 were included in the analysis. The overall or age-grouped distributions of age and thyroid lobe measurement among the males and females (table 1) did not show significant difference. Analysis for the subgroup NSG (Table 2) showed significant difference of AP diameter and volumes with the right lobe being significantly larger than the left lobe. Further analysis within groups shows non-retention of this difference for the group 1 (p > 0.05) meaning the pre-adolescent in this study had similar size lobes. The retention of difference in the group 2 (p > 0.05) indicates the finding of disparity of size between the thyroid lobes may be restricted for adolescent and adults. There was a weak correlation between age and total thyroid volume which was positive for all NGS patients and negative for NGS patients belonging to group 2, both without reaching a statistical significance (Figure 1 A and B). A significantly strong correlation was found between age and total thyroid volume for the NGS patients belonging to group 1 (Figure 1 C).

**Table 1:** Overall distribution of age and thyroid lobe measurements by gender.

|  | Male | Female | t-test p-value |
| --- | --- | --- | --- |
| All patients (n) | 20 | 90 |  |
| Age (year) | 27.8 ± 15.6 | 28.2 ± 12.7 | 0.92 |
| Right lobe AP (mm) | 16.1 ± 5.6 | 15.5 ± 4.2 | 0.66 |
| Right lobe volume (cc) | 7.7 ± 6.9 | 7.3 ± 5.8 | 0.82 |
| Left lobe AP (mm) | 14.4 ± 4.6 | 14.0 ± 4.9 | 0.73 |
| Left lobe volume (cc) | 6.2 ± 6.6 | 5.8 ± 7.0 | 0.79 |
| Total gland volume | 13.9 ± 13.3 | 13.1 ± 12.2 | 0.80 |
| Group 1 with age < 15 years (n) | 5 | 14 |  |
| Age (year) | 8.4 ± 3.7 | 9.7 ± 3.5 | 0.50 |
| Right lobe AP (mm) | 11.3 ± 2.6 | 14.0 ± 3.5 | 0.09 |
| Right lobe volume (cc) | 3.1 ± 1.9 | 5.2 ± 4.5 | 0.18 |
| Left lobe AP (mm) | 10.7 ± 3.4 | 12.3 ± 3.3 | 0.38 |
| Left lobe volume (cc) | 2.7 ± 2.0 | 3.9 ± 3.2 | 0.37 |
| Total gland volume | 5.8 ± 3.9 | 9.0 ± 7.7 | 0.25 |
| Group 2 with age ≥ 15 years (n) | 15 | 76 |  |
| Age (year) | 34.3 ± 12.1 | 31.6 ± 10.6 | 0.43 |
| Right lobe AP (mm) | 17.7 ± 5.5 | 15.7 ± 4.3 | 0.22 |
| Right lobe volume (cc) | 9.2 ± 7.4 | 7.7 ± 5.9 | 0.46 |
| Left lobe AP (mm) | 15.6 ± 4.4 | 14.3 ± 5.2 | 0.31 |
| Left lobe volume (cc) | 7.4 ± 7.2 | 6.1 ± 7.5 | 0.55 |
| Total gland volume | 16.6 ± 14.2 | 13.8 ± 12.8 | 0.49 |

**Table 2:** Thyroid gland measurements from normal sized glands (NSG) patients and the t-test p-values for the difference of means.

|  |  | Right lobe | Left lobe | t-test p-value |
| --- | --- | --- | --- | --- |
| NSG all (n=31) | AP diameter (mm) | 12.4 ± 1.4 | 11.3 ± 1.9 | 0.017* |
|  | Volume (cc) | 3.9 ± 1.4 | 3.1 ± 1.4 | 0.026* |
|  | Total gland volume | 6.9 ± 2.5 |  |  |
| NSG Group 1 (n=7) | AP diameter (mm) | 11.6 ± 1.9 | 10.2 ± 2.3 | 0.247 |
|  | Volume (cc) | 2.8 ± 1.5 | 2.0 ± 1.1 | 0.314 |
|  | Total gland volume | 4.8 ± 2.5 |  |  |
| NSG Group 2 (n=24) | AP diameter (mm) | 12.6 ± 1.2 | 11.6 ± 1.7 | 0.029* |
|  | Volume (cc) | 4.2 ± 1.2 | 3.4 ± 1.4 | 0.030* |
|  | Total gland volume | 7.6 ± 2.1 |  |  |

**Figure 1:**
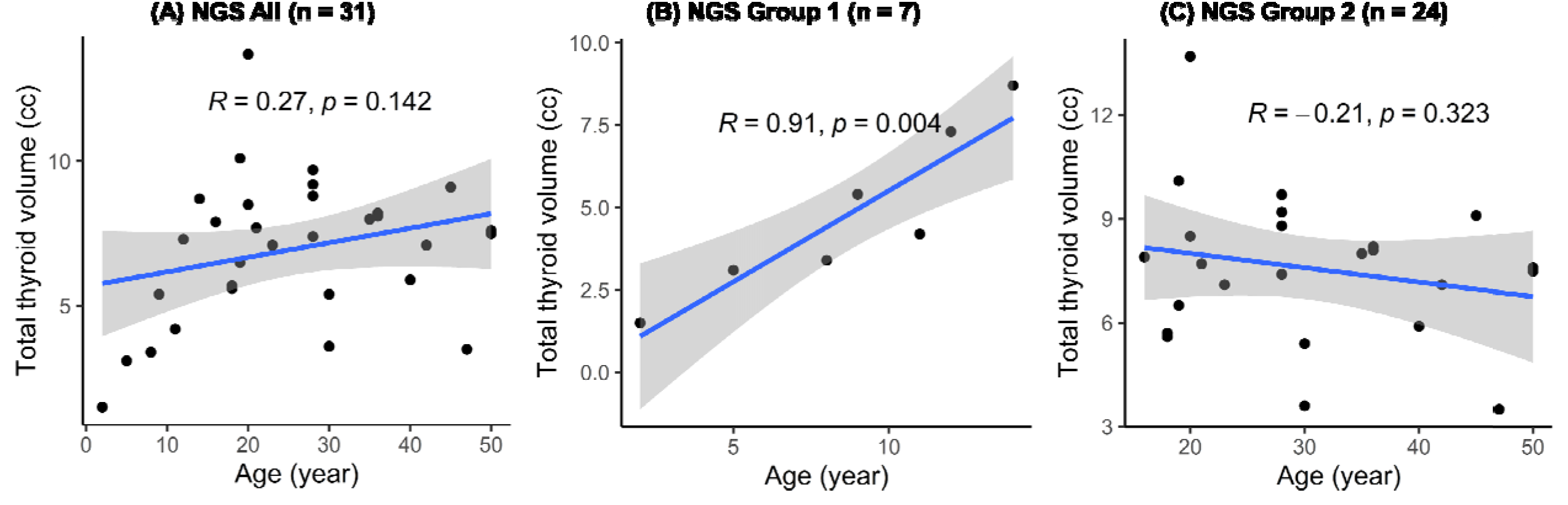
Scatter plots showing correlation of total thyroid gland volume (cc) in Y-axis with age (year) in X-axis from NGS all patients (A), NGS group 1 (B), and NGS Group 2 (C). The correlation co-efficient R and Pearson’s p-values are indicated within each plot.

According to figure 2, the mean volumes of the right lobe were slightly larger than those of left lobe in the categories of ‘normal’ and ‘moderate thyromegaly’. On the contrary, the mean volumes of the right lobe were slightly smaller than those of left lobe in the categories of ‘mild thyromegaly’ and ‘marked thyromegaly’. According to figure 3, the AP diameter for left lobe (in comparison to that for right lobe), when used to classify the total gland volumes, the number of patients belonging to normal volume was higher with a slightly higher mean of total gland volume. Consequently, the number of patients belonging to enlarged categories were lower, however, with higher mean of total gland volume. Based on these findings, it may be permitted to assume that the when the AP diameter categories of right lobe was used the normal glands were better classified whereas the AP diameter categories of left lobe allowed a better classification of the enlarged glands. This indicates the possibility of conditions with unilateral thyroid lobe enlargement with the total gland volume remaining normal. The forest plot (figure 4) is a visual representation of the sprawling normal references ranges for the AP dimeter of thyroid lobe which can be compared in the same plot with the reported values of the current study.

**Figure 2:**
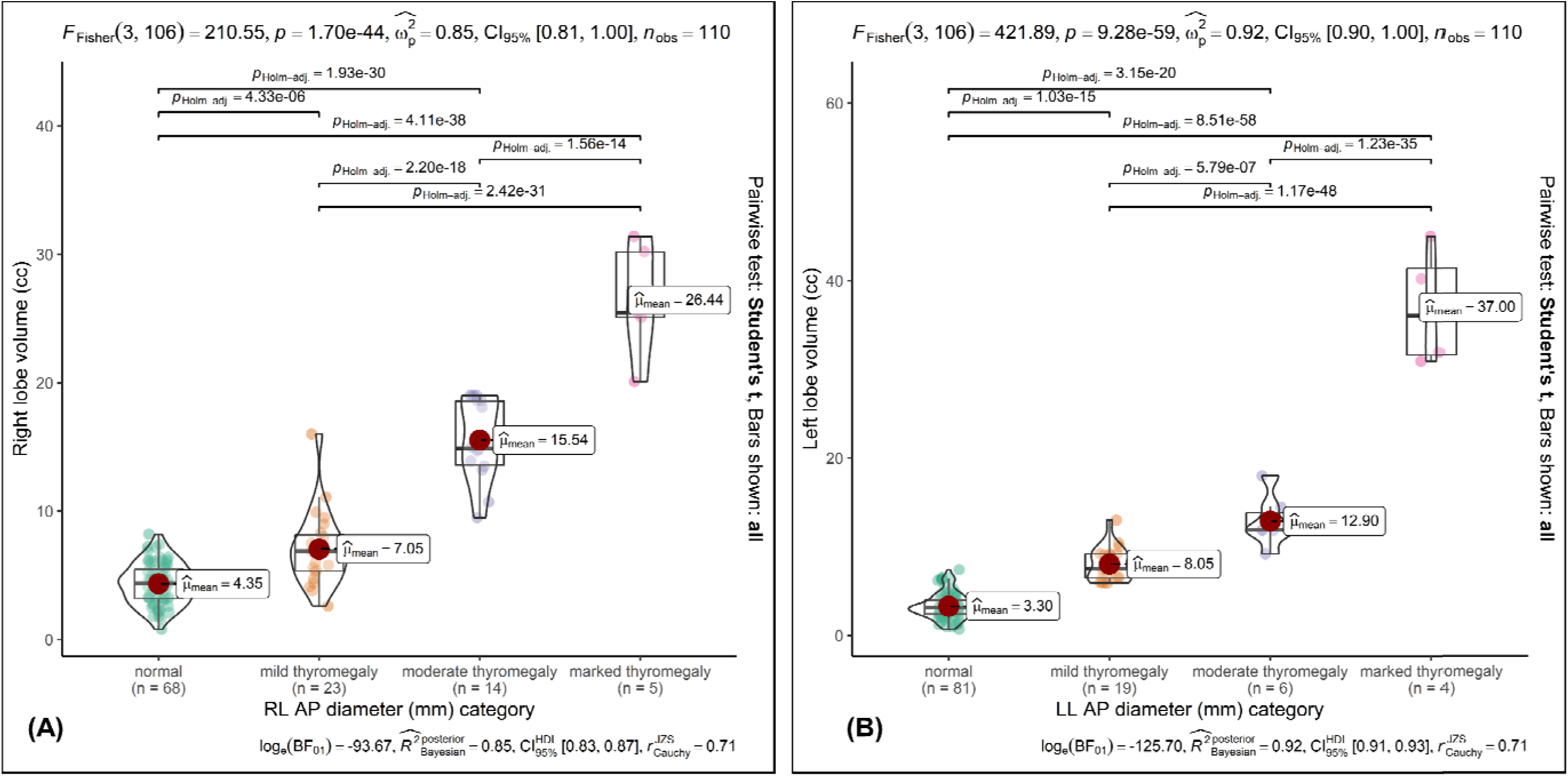
Box-violin plots showing distribution of right lobe volume across the arbitrary AP diameter categories (A), and left lobe volume across the arbitrary AP diameter categories (B). The significance of pair-wise difference is indicated at the top bars.

**Figure 3:**
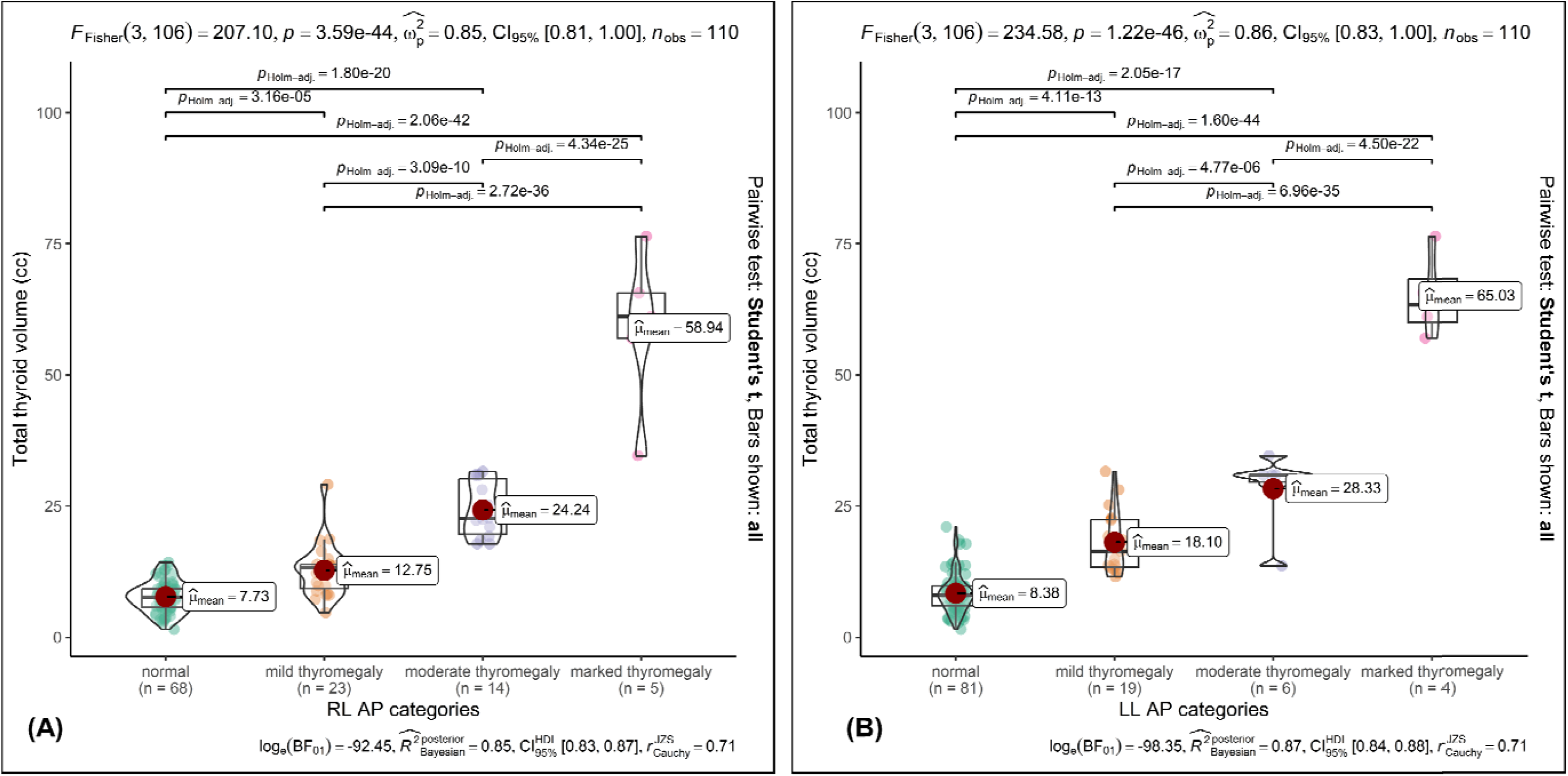
Box-violin plots showing distribution of total gland volume across the arbitrary AP diameter categories for the right lobe (A), and the same for the left lobe (B). The significance of pair-wise difference is indicated at the top bars.

**Figure 4:**
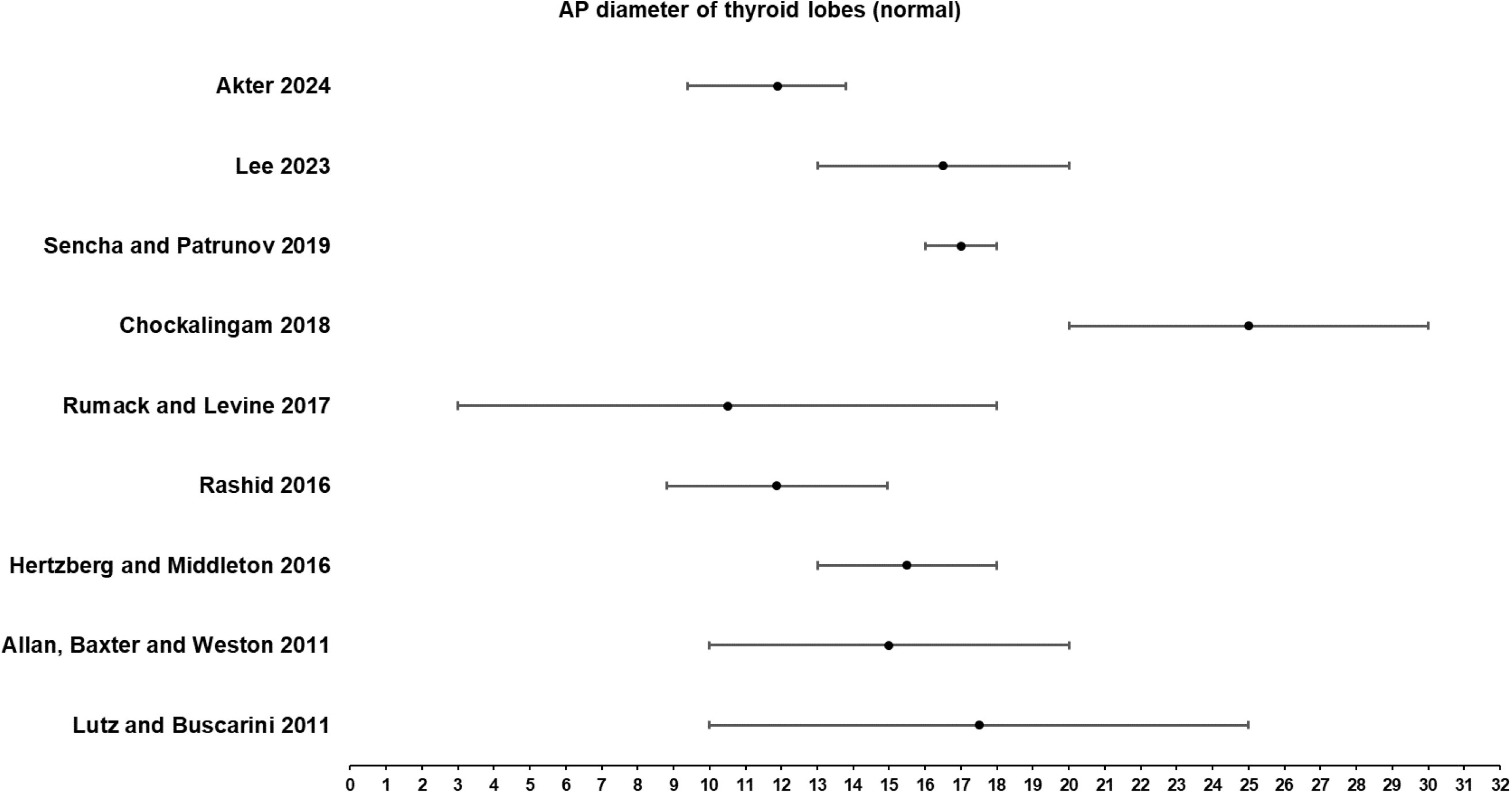
Forest plots showing reported normal ranges and mean values for AP diameter of thyroid lobe from several references along with the values observed in the current study.

## DISCUSSION

Larger proportion of the patients in this study were females who in the group 1 tended to be older than males and tended to have larger thyroids, while the females in the group 2 tended to be younger than males and tended to have smaller thyroids, although without reaching a significance. The thyroid size in males is reportedly larger than the age matched females, and there is an observed increment of thyroid size with the progress of age (8, 18-25, 28, 31, 35, 38, 39). Thus, the larger thyroid size in females of group 1 is due to their older age who tended to have larger thyroid and the finding in group 2 almost matches with the reported results, where the small sample size precluded the attainment of a statistical significance.

An age and sex matched comparison of thyroid sizes could be done in this study due to the small sample size. It is known that the age and sex related difference is not simple because of thyroid size may increase due to pregnancy, obesity in females, consumption of oral contraceptive in females, cigarette smoking, exposure to environmental pollutant and pesticides, alcohol consumption and winter season and diabetes while a decrease of thyroid size may occur in chronic renal disease, chronic alcoholism and acute hepatic diseases (26, 30, 32-34). Assessing the contribution of these factors to thyroid gland size was beyond the scope of the current study.

The finding of right lobe being larger than the left in NSG patients in the current study, matches with a recent series (38). The finding of significant positive correlation in the NSG group 1 consisting of pre-adolescents in this study matches with a reported fact (37) that correlation between thyroid volume and age is stronger before the age of 18. In the NSG group 2 consisting of 19 females and five males there is a weak negative correlation of thyroid gland volume with that of age although without reaching significance. Similar negative correlation of thyroid size and age in females were reported by a French study (27). This study shows a right-left disparity of lobe enlargement across the enlargement categories, and it is possible to have normal total thyroid volume with single lobe enlarged. Therefore, interpretation of thyroid lobe volume should always be accompanied by interpretation of individual lobes’ volume.

Currently, the non-standard arbitrary range for AP diameter of thyroid lobes does not have a lower margin to classify the small sized thyroid gland. As a result, the observation of thyroid volumes across the arbitrary AP diameter ranges remains inconclusive about the small volume thyroid while at the same time there remains a high possibility that some small volume thyroid glands may have been considered within the normal volume glands. Specific observation of thyroid glands’ size in patients with pituitary or hypothalamic hypothyroidism or in patients with initially normal sized glands requiring thyroxine replacement therapy, may fill the current void of a standard for small volume thyroid gland.

## CONCLUSION

This study describes the relationship of the two demographic traits, age and sex with thyroid size in small series of patient. Moreover, this study shows significant asymmetry between the thyroid lobes which was likely responsible for the disproportionate distribution of thyroid volume across the arbitrary AP diameter categories of thyroid lobes. Thus, it is important to consider the natural asymmetry between the thyroid lobes for accurate classification of thyroid volumes. This study points toward the void for a reference local standard of thyroid volume that can classify not only the different categories of enlargement but also the small volume glands.

## Data Availability

All data produced in the present study are available upon reasonable request to the corresponding author

## Notes

### Competing Interest Statement

The authors have declared no competing interest.

### Funding Statement

This study did not receive any funding

### Author Declarations

IRB of Institute of Nuclear Medicine and Allied Sciences, Gopalganj waived ethical approval for this work

